# Judgements about carer assessments for carers of people with dementia: case vignette study

**DOI:** 10.1101/2024.04.11.24305691

**Authors:** Paul Clarkson, Lucie Mills, Asif Butt, Baber Malik, Ruth Eley, Cecilia Toole, Caroline Sanders, Ian Sheriff

## Abstract

**Objectives:** UK carer assessments, in primary and social care, intend to discover what carers need in their caring roles and more widely. Evidence points to these not being configured sufficiently around carers of people with dementia, with potentially their breadth of needs not being recognised. We evaluated the extent of agreement, between carers of people with dementia, primary care, and social care professionals, on their recommendations from assessing carers’ needs in a range of circumstances. It is intended for findings to be taken forward as recommendations for policy and practice.

**Methods:** Comparison of judgements, between carers, primary and social care professionals, on whether real-life circumstances in 9 anonymised case vignettes necessitated a range of 14 services to support carers appropriately. Participants were 6 carers of people with dementia, 7 primary care staff, and 2 social care staff. We presented participants with each vignette and asked them to make binary judgements of whether they would recommend a range of services in each case. Percentage agreement and Fleiss’ kappa coefficients measured the level of agreement amongst multiple carers, primary and social care staff and overall. These agreements were then compared.

**Results:** Carers agreed in their judgements more than primary or social care professionals. The overall level of agreement from judgements made by all participants, however, was ‘slight’ with variability between participant groups and overall. The need for First Language Support in some cases was recognised, an improvement from previous evidence.

**Conclusions:** Case vignettes are useful for investigating judgements concerning these carers’ needs, so raising issues for policy and practice. It is essential for carer assessments to be more reliable in recommending services based on need to ensure less variability, depending on assessor and carers circumstances.

## Introduction

Over 670,000 people (UK) are primary, unpaid carers for people with dementia [1]. Carer assessments are a formal mechanism for identifying what carers need and require in their caring roles and more widely. In social care, these assessments are a legal duty, [2] and in primary care are tied to incentives and are more discretionary, often linked to health checks and signposting by general practices [3].

These assessments are not specific to carers of people with dementia, although their early effects for this group of carers, before current legislation under the 2014 Care Act, were evidenced [4]. Evidence points to these assessments not being configured sufficiently around carers of people with dementia, with potentially their breadth of needs not being recognised. More recent evidence points to these carers often not receiving an assessment, despite this being mandated, with often carers not seeing these assessments as valuable [5]. Although carer assessments are a requirement, their content and the types of needs judged important for those carers of people with dementia are variable and often not recognised.

The experiences of two groups of carers of people with dementia are of note. Those carers from south Asian backgrounds report that the multiplicity of informal carer networks and the distinct needs of second-generation carers are not recognised or neglected [6]. Carers of people living with young onset dementia have distinct needs, which also tend not to be recognised, including impact on relationships, financial implications, and a sense of loss [7].

As part of a wider study, we aimed to examine whether the needs and preferences of carers of people with dementia were potentially covered sufficiently in these assessments. We aimed to evaluate the extent of agreement, between carers of people with dementia, primary care, and social care professionals, on their recommendations from assessing carers’ needs in a range of circumstances. We used case vignettes, relatively short accounts describing realistic carer’s situations, as a method to identify how different groups of raters assessed identical scenarios.

## Material and Methods

### Design

Nine detailed case vignettes of anonymised real-life cases of carers’ circumstances were designed and used. These vignettes were created to express a range of situations that carers may be in as they interfaced with health and social care systems. The vignettes were given to participants belonging to three groups: carers of people with dementia, primary care staff, and social care staff. Participants were approached, respectively, through national and local carers’ charities, general practice and Primary Care Network, and local authority consortium. All participants gave informed consent to take part. Each participant was told that the vignettes were based on real cases and experiences but were anonymised as to person and place. Vignettes were completed online, through Zoom, email, or (for primary care staff) in person at a workshop meeting. Each participant was asked to read each vignette and make a binary judgement (yes/no) of whether they would recommend a range of services (n=14) in each case, that were warranted to help support the carers’ needs. Participants were also given the option of describing their reasons behind the judgements. The vignettes and responses were returned anonymously to the research team.

### Case vignettes

This was a co-produced study with carers and carer representative organisations. The anonymised vignettes were written jointly by carers of people with dementia, through carers organisations and members of the research team, taken from carers’ experiences and circumstances. The vignettes were designed to express a range of different circumstances and possible service responses, but particularly (although not exclusively) focusing on the needs of carers of people with younger onset dementia and those from a south Asian background. The potential needs of carers from these groups, highlighted in vignettes, included financial, employment and relationship need, [8] multigenerational caring, language, and cultural needs [9]. Possible service responses to the vignettes, which participants were asked to indicate if warranted, were included after consultation with carers groups and charities supporting carers of people with dementia. Fig 1 shows an example of an anonymised case vignette with the same standardized format used for all vignettes.

**Fig 1.**
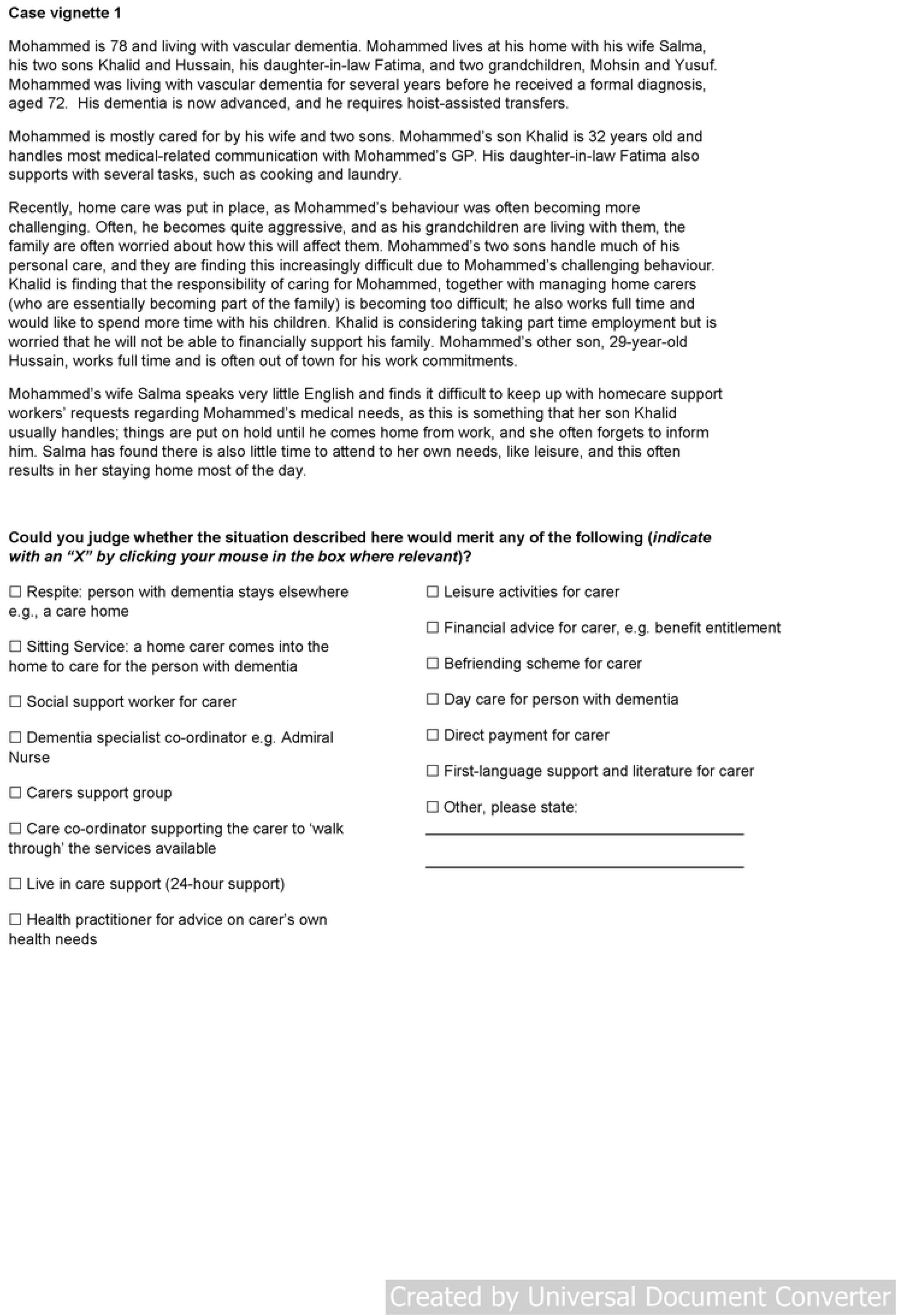
Sample vignette. Description of carer’s circumstances and request to signal service responses that would be considered appropriate.

Each vignette expressed circumstances, presenting insights from carers’ perspectives to generate participants’ judgements, but without leading too directly. The vignettes had an average length of 180 words (ranging from 91 to 318). In general, each vignette took participants about 5-10 minutes to read and complete.

### Participants

All participants were approached in writing with Participant Information Sheets and invited to take part in the study and had to give their written consent to take part. Recruitment took place between 14/12/2022 and 31/12/2023. Carers of people with dementia were interested carers who had received carers assessments before and who wished to express their views about carers’ needs in the context of these assessments. These carers were not involved in the construction of the case vignettes. Primary care staff were professionals working in or with primary care practices who had an interest in dementia or older people’s healthcare. Social care staff were present or former statutory assessors within the context of carer assessments and were social workers of local authorities. It was left to participants to decide which of the nine vignettes they rated and the completion of reasons for their judgments was entirely optional.

### Ethics statement

The study was approved by the University of Manchester Research Ethics Committee (Ref: 2022-14569-26301; 14/12/2022) and, additionally, for primary care staff, the Health Research Authority (IRAS Project ID: 326181; 23/HRA/1321).

### Statistical analysis

Data analysis was undertaken using SPSS v28 for Windows. Descriptive statistics (percentage agreement) were used to compare judgements of each of the groups, about whether each of the possible 14 service responses was warranted in each case. Fleiss’ kappa coefficients, for multiple raters, [10] measured the level of agreement (inter-rater reliability) amongst carers, primary and social care staff and overall. the Kappa coefficient. Based on standards outlined by Landis and Koch [11], a Kappa coefficient <0 was considered to be poor agreement, 0–0.20 slight, 0.21–0.40 fair, 0.41–0.60, moderate, 0.61–0.80 substantial, and 0.81–1.0 almost perfect agreement. Tests were two-tailed and P < 0.05 was considered statistically significant.

## Results

Full data were available from 6 carers, 7 primary care staff and 2 social care staff. Two carers cared for people with younger onset dementia. Primary care staff included general practitioners, an assistant practice manager, nurses, a practice pharmacist, and a paramedic. Social care staff were social workers familiar with statutory carer’s assessments.

### Comparison of judgements by carers and professional staff

Table 1 summarises the agreement of participants in each group and overall concerning whether they would recommend each of the 14 services in each case. Percentage agreement and Fleiss’ kappa coefficients are compared. Total agreement (100%) between all groups was seen in only two cases: case vignette 8 for Sitting Service and case vignette 1 for Financial Advice. There was relatively high agreement for case vignette 7, Sitting Service; case vignette 8 for Financial Advice; and case vignette 1, First Language Support, which was a case of a carer and person with dementia from a south Asian background. The remaining cases showed a range of differing judgements about whether each service was recommended.

**Table 1.** Proportion (%) of each group judging case warrants service and overall level of agreement; Fleiss’ kappa (95% confidence interval); p value, between participants.

| Case vignette | carers | Primary care staff | Social care staff | Total |
| --- | --- | --- | --- | --- |
| ***Respite care for person with dementia*** |  |  |  |  |
| 1 | 67 | 57 | 100 | 67 |
| 2 | 17 | 43 | 0 | 27 |
| 3 | 17 | 29 | 50 | 27 |
| 4 | 83 | 29 | 100 | 60 |
| 5 | 67 | 14 | 50 | 40 |
| 6 | 33 | 43 | 50 | 33 |
| 7 | 33 | 43 | 50 | 40 |
| 8 | 67 | 57 | 100 | 67 |
| 9 | 83 | 100 | 100 | 93 |
| Fleiss' kappa | 0.11 (-0.09 to 0.28);<br>p=0.20 | 0.08 (-0.06 to 0.23);<br>p=0.25 | 0.30 (-0.35 to 0.95);<br>p=0.37 | 0.12 (0.06 to 0.18);<br>**p<0.001** |
| ***Sitting service*** |  |  |  |  |
| 1 | 83 | 71 | 100 | 87 |
| 2 | 33 | 29 | 50 | 33 |
| 3 | 50 | 57 | 50 | 53 |
| 4 | 83 | 86 | 50 | 80 |
| 5 | 17 | 29 | 50 | 27 |
| 6 | 33 | 43 | 50 | 33 |
| 7 | 100 | 100 | 50 | 93 |
| 8 | 100 | 100 | 100 | 100 |
| 9 | 50 | 57 | 50 | 53 |
| Fleiss' kappa | 0.24 (0.07 to 0.40);<br>**p=0.006** | 0.18 (0.04 to 0.32);<br>**p=0.01** | -0.35 (-1.00 to 0.15);<br>p=0.30 | 0.12 (0.17 to 0.30);<br>**p<0.001** |
| ***Social support worker for carer*** |  |  |  |  |
| 1 | 100 | 86 | 50 | 87 |
| 2 | 67 | 86 | 0 | 67 |
| 3 | 83 | 86 | 0 | 73 |
| 4 | 83 | 71 | 0 | 67 |
| 5 | 83 | 71 | 0 | 67 |
| 6 | 33 | 86 | 0 | 53 |
| 7 | 83 | 86 | 0 | 73 |
| 8 | 83 | 86 | 0 | 73 |
| 9 | 100 | 86 | 100 | 93 |
| Fleiss' kappa | 0.06 (-0.10 to 0.23);<br>p=0.46 | -0.14 (-0.28 to 0.004); p=0.06 | 0.60 (-0.05 to 1.25);<br>p=0.07 | -0.005 (-0.07 to 0.06); p=0.87 |
| ***Dementia specialist coordinator, e.g. Admiral Nurse*** |  |  |  |  |
| 1 | 100 | 71 | 100 | 87 |
| 2 | 0 | 14 | 50 | 13 |
| 3 | 0 | 14 | 50 | 13 |
| 4 | 83 | 43 | 100 | 67 |
| 5 | 17 | 43 | 50 | 33 |
| 6 | 17 | 57 | 50 | 40 |
| 7 | 33 | 71 | 50 | 53 |
| 8 | 67 | 71 | 50 | 67 |
| 9 | 83 | 57 | 100 | 73 |
| Fleiss' kappa | 0.46 (0.29 to 0.63);<br>**p<0.001** | 0.05 (-0.09 to 0.19); p=0.52 | -0.50 (-1.15 to 0.15); p=0.13 | 0.19 (0.13 to 0.25);<br>**p<0.001** |
| ***Carers support group*** |  |  |  |  |
| 1 | 100 | 67 | 100 | 80 |
| 2 | 50 | 86 | 100 | 73 |
| 3 | 100 | 43 | 50 | 67 |
| 4 | 67 | 71 | 100 | 73 |
| 5 | 83 | 43 | 50 | 60 |
| 6 | 33 | 43 | 0 | 33 |
| 7 | 100 | 57 | 50 | 73 |
| 8 | 83 | 57 | 100 | 73 |
| 9 | 100 | 71 | 50 | 80 |
| Fleiss' kappa | 0.20 (0.03 to 0.37);<br>**p=0.02** | -0.07 (-0.21 to 0.07); p=0.34 | 0.00 (-0.65 to 0.65);<br>p=1.00 | 0.02 (-0.04 to 0.08);<br>p=0.54 |
| ***Care coordinator, supporting carer to 'walk through' services available*** |  |  |  |  |
| 1 | 67 | 71 | 50 | 67 |
| 2 | 83 | 71 | 100 | 80 |
| 3 | 83 | 71 | 100 | 80 |
| 4 | 100 | 57 | 100 | 80 |
| 5 | 83 | 86 | 100 | 87 |
| 6 | 50 | 86 | 50 | 67 |
| 7 | 83 | 71 | 100 | 80 |
| 8 | 83 | 86 | 100 | 87 |
| 9 | 83 | 86 | 100 | 87 |
| Fleiss' kappa | -0.07 (-0.24 to 0.09); p=0.39 | -0.11 (-0.25 to 0.03); p=0.14 | -0.12 (-0.78 to 0.53); p=0.71 | -0.04 (-0.10 to 0.03); p=0.26 |
| ***Live in care support (24-hour support)*** |  |  |  |  |
| 1 | 0 | 57 | 0 | 33 |
| 2 | 17 | 28 | 0 | 20 |
| 3 | 50 | 14 | 0 | 20 |
| 4 | 17 | 14 | 0 | 13 |
| 5 | 17 | 28 | 0 | 20 |
| 6 | 33 | 14 | 0 | 20 |
| 7 | 0 | 28 | 0 | 13 |
| 8 | 17 | 28 | 0 | 20 |
| 9 | 50 | 57 | 0 | 47 |
| Fleiss' kappa | -0.03 (-0.20 to 0.14); p=0.75 | -0.03 (-0.17 to 0.11); p=0.68 | N/A service not recommended in all cases | -0.02 (-0.08 to 0.05); p=0.62 |
| **Health practitioner, advice on carer's own health needs** |  |  |  |  |
| 1 | 67 | 57 | 100 | 67 |
| 2 | 17 | 57 | 0 | 33 |
| 3 | 0 | 29 | 0 | 13 |
| 4 | 17 | 43 | 0 | 27 |
| 5 | 0 | 29 | 50 | 20 |
| 6 | 0 | 0 | 0 | 0 |
| 7 | 33 | 43 | 0 | 33 |
| 8 | 0 | 57 | 0 | 27 |
| 9 | 17 | 86 | 50 | 53 |
| Fleiss' kappa | 0.17 (0.005 to 0.34); **p=0.04** | 0.07 (-0.06 to 0.22); p=0.28 | 0.36 (-0.30 to 1.01); p=0.28 | 0.11 (0.05 to 0.17); **p<0.001** |
| **Leisure activities for carer** |  |  |  |  |
| 1 | 67 | 28 | 50 | 47 |
| 2 | 17 | 28 | 0 | 27 |
| 3 | 33 | 28 | 50 | 33 |
| 4 | 33 | 28 | 0 | 27 |
| 5 | 33 | 28 | 0 | 27 |
| 6 | 0 | 14 | 0 | 7 |
| 7 | 33 | 57 | 50 | 47 |
| 8 | 50 | 86 | 100 | 73 |
| 9 | 83 | 57 | 50 | 67 |
| Fleiss' kappa | 0.05 (-0.12 to 0.22); p=0.57 | 0.05 (-0.09 to 0.19); p=0.50 | 0.00 (-0.65 to 0.65); p=1.00 | 0.11 (0.04 to 0.17); **p<0.001** |
| **Financial advice for carer, e.g., benefit entitlement** |  |  |  |  |
| 1 | 100 | 100 | 100 | 100 |
| 2 | 100 | 86 | 100 | 87 |
| 3 | 67 | 57 | 100 | 67 |
| 4 | 67 | 57 | 100 | 60 |
| 5 | 67 | 57 | 50 | 60 |
| 6 | 67 | 57 | 50 | 53 |
| 7 | 83 | 86 | 100 | 87 |
| 8 | 100 | 43 | 100 | 73 |
| 9 | 100 | 86 | 100 | 93 |
| Fleiss' kappa | 0.01 (-0.12 to 0.18); p=0.88 | 0.02 (-0.15 to 0.16); p=0.78 | -0.12 (-0.78 to 0.53); p=0.71 | 0.06 (0.00 to 0.12); **p=0.05** |
| **Befriending scheme for carer** |  |  |  |  |
| 1 | 50 | 43 | 50 | 47 |
| 2 | 17 | 28 | 0 | 20 |
| 3 | 17 | 28 | 0 | 20 |
| 4 | 17 | 14 | 0 | 13 |
| 5 | 0 | 28 | 0 | 13 |
| 6 | 0 | 43 | 0 | 20 |
| 7 | 17 | 43 | 0 | 27 |
| 8 | 33 | 57 | 50 | 47 |
| 9 | 33 | 57 | 50 | 47 |
| Fleiss' kappa | -0.03 (-0.19 to 0.14); p=0.71 | -0.07 (-0.21 to 0.07); p=0.29 | -0.20 (-0.03 to 0.45); P=0.55 | 0.03 (-0.03 to 0.09); P=0.41 |
| **Day care for person with dementia** |  |  |  |  |
| 1 | 67 | 57 | 50 | 60 |
| 2 | 17 | 57 | 50 | 40 |
| 3 | 0 | 43 | 50 | 20 |
| 4 | 67 | 43 | 100 | 60 |
| 5 | 33 | 14 | 50 | 27 |
| 6 | 17 | 29 | 50 | 27 |
| 7 | 50 | 100 | 50 | 73 |
| 8 | 67 | 71 | 100 | 73 |
| 9 | 67 | 86 | 100 | 80 |
| Fleiss' kappa | 0.11 (-0.06 to 0.27);<br>p=0.22 | 0.14 (0.00 to 0.28); **p=0.05** | -0.50 (-1.15 to 0.15); p=0.13 | 0.11 (0.05 to 0.18);<br>**p<0.001** |
| **Direct payment for carer** |  |  |  |  |
| 1 | 50 | 71 | 100 | 67 |
| 2 | 17 | 43 | 50 | 33 |
| 3 | 50 | 43 | 50 | 47 |
| 4 | 50 | 29 | 0 | 33 |
| 5 | 50 | 57 | 50 | 53 |
| 6 | 0 | 29 | 0 | 13 |
| 7 | 50 | 86 | 50 | 67 |
| 8 | 17 | 17 | 0 | 13 |
| 9 | 83 | 57 | 50 | 67 |
| Fleiss' kappa | 0.08 (-0.08 to 0.24);<br>p=0.35 | 0.04 (-0.09 to 0.19); p=0.53 | -0.17 (-0.82 to 0.48); p=0.61 | 0.11 (0.05 to 0.17);<br>**p<0.001** |
| **First language support, literature for carer** |  |  |  |  |
| 1 | 100 | 57 | 100 | 80 |
| 2 | 0 | 14 | 0 | 6 |
| 3 | 0 | 28 | 0 | 13 |
| 4 | 0 | 28 | 0 | 13 |
| 5 | 0 | 28 | 0 | 13 |
| 6 | 0 | 0 | 0 | 0 |
| 7 | 0 | 14 | 0 | 6 |
| 8 | 0 | 0 | 0 | 0 |
| 9 | 0 | 14 | 0 | 6 |
| Fleiss' kappa | 1.00 (0.83 to 1.17);<br>**p<0.001** | 0.03 (-0.11 to 0.17); p=0.67 | 1.00 (0.34 to 1.65);<br>**p=0.003** | 0.37 (0.31 to 0.43);<br>**p<0.001** |

Vignettes 1,3,4, and 5 = carers of people with dementia from south Asian background; vignettes 2 and 7 = carers of people with young onset dementia

Kappa values for all participant groups showed only ‘slight’ agreement [11] across the case vignettes, but ‘fair’ agreement for First Language Support. Across the groups (Total column in Tables 1 and 2), agreement for Social Support Worker, Care Co-ordinator and Live in Carer was no better than chance. Carers had the highest level of agreement: ‘slight’ (9 services), ‘moderate’ (1 service) and ‘almost perfect’ (1 service; First Language Support). For carers, agreement on three services (Care Co-ordinator, Live in Care, Befriending Scheme) was no better than chance. Primary care staff had ‘slight’ agreement for 8 services, ‘fair’ agreement for 1 service, with agreement no better than chance for the remaining 5 services. Social care staff had the lowest level of agreement, ‘slight’ (2 services) fair’ (2 services), ‘moderate (1 service), and no better than chance for the remaining 8 services. However, social care staff had ‘almost perfect’ agreement on the need for First Language Support.

### Reasons behind the judgements

Examination of the stated reasons behind the judgements made by participants in the vignettes helped to discern some of the wider factors influencing decision-making in these assessments. These tended to vary according to the groups of participants undertaking the exercise. For carers, judgements were made based on perceived risk, such that if there was a notion that needs were increasing to an unsustainable level then more and different services would be recommended.

Carers often recommended services with the aim of building confidence in the carer’s ability to continue caring. Carers were also concerned about people’s entitlements and ensuring that these were supported through assessments but also that this depended on the local supply of services where the carer lived. Primary care staff, perhaps understandably, stressed the need for a formal dementia diagnosis for the carer’s relative, so that they could be keyed into the support services available. The thought of ‘carer burnout’ was also evident in the reasoning of primary care staff and that the carers needed to know all the options and support available to them to perhaps prevent that. Social care staff again stressed a consideration with carers’ entitlements and ensuring that they were eligible for certain services and financial help. Assessment was not seen as a one-off process by social care staff, and there was a notion of establishing a continuity to decisions and referring the carer on further for help from other professionals.

## Discussion

The nature and process of carer assessments are no different than for assessments in general. Previous work on social care assessments has shown wide variability, across professionals, in the identification of older people’s needs [12,13]. For carer assessments of those caring for someone with dementia, this variability continues to be an issue, with many carers dissatisfied that their needs had not been identified or met [14,15]. In this case vignette study, we found wide variability in the reliability to which carers’ needs and suitable service responses were identified that concur with some of these judgements. Total agreement on needs occurred in only two vignettes for two service responses and there was mostly only slight agreement across the case vignettes. Carers tended to agree the most, with primary care staff second and social care staff agreeing the least.

There was relatively high agreement in terms of the need for First Language Support, amongst carers and social care staff (showing ‘almost perfect’ agreement), particularly for case vignette 1 which was a case of a south Asian carer. This finding is encouraging considering previous evidence of this type of support being undervalued in carer assessments and support [16]. On the other hand, there was relatively low agreement by professionals on the need for Befriending Support, for the vignettes focusing on south Asian carers. This type of support has been highly valued by south Asian carers [16] and is seen as providing a different focus than more traditional services such as respite and day care.

### Strengths and Limitations of the Study

Our study has limitations. We examined the judgements of a relatively small number of participants, [17]particularly social care staff. This limits the statistical power by which the indices of agreement were produced. Smaller samples also raise the potential of bias, in that it may be likely for raters who tended to choose particular categories (service responses) as present or absent to influence the results. This can produce situations where there is high percentage agreement amongst raters but a low kappa [18]. This was the case for our study, where, for example, for Financial Advice and Dementia Specialist Coordinator, rated by social care staff, there was high percentage agreement but with very low kappa scores, resultant agreement being no better than chance.

However, this notwithstanding, our study has some strengths. We used real-life, detailed vignette descriptions of carers’ circumstances and these were able to prompt responses that mimic the decision-making processes of assessments. This makes it possible to draw conclusions that have ecological validity [19] and can be more realistically generalised to inform practice and policy. There is no ‘gold standard’ by which to judge the reliability of assessments, particularly for assessments that have a legislative basis, such as these carer assessments in the UK. The assessment process is complex and variable and is influenced by many factors, both internal and external to the assessment ‘encounter’. In addition, there is very little prior research investigation of how carer assessments operate in primary care in the UK, primarily because they do not have a legislative basis and are informed only by guidance, usually as part of UK health checks. Thus, this study adds to the evidence base in this area by considering the judgements of social care and primary care staff against those of carers, the subject of these assessments. Carers, arguably, are the best judge of their own needs for support and it is noteworthy that carers here agreed the most across the service responses and vignettes. Our study design recognised this in terms of co-producing the vignettes with carers to stimulate participant’s judgement decisions.

## Conclusion

Case vignettes are a useful tool for investigating different professionals’ judgements concerning the needs of carers of people with dementia and so raising issues for policy and practice. This study showed these assessments can still be highly variable in terms of identifying the kinds of needs and service responses that might be required to support carers in their caring role. The need for First Language Support in some cases was recognised, an improvement from previous evidence.

However, efforts to improve the reliability with which carer assessments are undertaken, so that prominent needs are identified and acted on in a consistent way, are still required.

## Data Availability

The datasets generated and analysed in this study and the full anonymized case vignettes are publicly available via the Figshare data repository, at https://doi.org/10.48420/25483270.v1 Participants consented to make anonymized data publicly available.

## Acknowledgements

We thank the carers, primary care, and social care staff for their time in taking part in the study. Together in Dementia Everyday (TIDE), Sight and Mind CIC, Dementia United (Greater Manchester), Kirklees Council, Bury Primary Care Network, and the Greater Manchester Social Work Academy assisted in the recruitment of participants.

## References

1. Alzheimer’s Society. Alzheimer’s Society’s view on carer support. 2014; Available from: https://www.alzheimers.org.uk/about-us/policy-and-influencing/what-we-think/carer-support

2. Seddon D, Robinson C. Carer assessment: continuing tensions and dilemmas for social care practice. Health Soc Care Community. 2015; 23(1): 14–22.

3. BMA/NHS England. Quality and Outcomes Framework guidance for 2021/22. 2021. Available from: https://www.england.nhs.uk/wp-content/uploads/2021/03/B0456-update-on-quality-outcomes-framework-changes-for-21-22-.pdf

4. Seddon D, Robinson C. (2001) Carers of older people with dementia: assessment and the Carers Act. Health Soc Care Community. 2021; 9(3): 151–158.

5. Newbronner L, Chamberlain R, Borthwick R, Baxter M, Glendinning C. A Road Less Rocky – Supporting Carers of People with Dementia. London, Carer’s Trust. 2013 Available at: https://carers.org/resources/all-resources/84-a-road-less-rocky-a-supporting-carers-of-people-with-dementia

6. Giebel CM, Zubair M, Jolley D, Bhui KS, Purandare N, Worden A, et al. South Asian older adults with memory impairment: improving assessment and access to dementia care. Int J Geriatr Psychiatry. 2015; 30(4): 345–56.

7. Together In Dementia Everyday (TIDE). Young Onset Dementia Survey Findings. 2020. Available from: https://www.tide.uk.net/young-onset-dementia/

8. Stamou V, La Fontaine J, Gage H, Jones B, Williams P, O’Malley M, et al. Services for people with young onset dementia: The ‘Angela’ project national UK survey of service use and satisfaction. Int J Geriatr Psychiatry. 2021; 36: 411–422.

9. Baghirathan S, Cheston R, Hui R, Chacon A, Shears P, Currie K. A grounded theory analysis of the experiences of carers for people living with dementia from three BAME communities: Balancing the need for support against fears of being diminished. Dementia. 2020; 19(5); 1672–1691.

10. Fleiss JL. Measuring nominal scale agreement among many raters. Psychol Bull. 1971; 76: 378–82.

11. Landis JR, Koch GG. The measurement of observer agreement for categorical data. Biometrics. 1977; 33: 159–74.

12. Clarkson P. What research tells social workers about their work with older people. In: Davies M, editor. Social Work with Adults. Basingstoke, Hampshire: Palgrave Macmillan; 2012. pp. 300–313.

13. Ellis K. Squaring the circle: user and carer participation in needs assessment. York: Joseph Rowntree Foundation; 1993.

14. Carers UK. (2022) State of Caring 2022 report. London: carers UK. 2022. Available from: https://www.carersuk.org/media/p4kblx5n/cukstateofcaring2022report.pdf

15. Stock C, Lambert S. (2011) Who cares wins? Carers’ experiences of assessment since the introduction of the Carers (Equal Opportunities) Act 2004. Research Policy and Planning. 2011; 28: 173–184.

16. Hepworth D. (2005) Asian carers’ perceptions of care assessment and support in the community. Br J Soc Work. 2005; 35: 337–353.

17. Sim J, Wright CC. The kappa statistic in reliability studies: use, interpretation, and sample size requirements. Phys Ther. 2005; 85: 257–68.

18. Dettori JR, Norvell DC. Kappa and beyond: Is there agreement? Global Spine J. 2020; 10(4): 499–501.

19. Kieffer S. ECOVAL: Ecological validity of cues and representative design in user experience evaluations. AIS Transactions on Human-Computer Interaction. 2017; 9 (2): 149–172.

